# Association of Digoxin Use at Norwood Discharge with Fontan Completion: A Study from the Pediatric Heart Network Public Dataset

**DOI:** 10.64898/2026.06.17.26355912

**Authors:** Alaa Aljiffry, Andrew Jergel, Yijin Xiang, Matthew E. Oster, Lazaros Kochilas

## Abstract

**Background:** Digoxin use after the Norwood procedure has been associated with improved interstage survival in hypoplastic left heart syndrome and related conditions. Whether this benefit translates into improved longer-term outcomes through staged palliation remains unknown. We aimed to determine the association of digoxin use at Norwood discharge with transplant-free survival and Fontan completion.

**Methods:** We conducted a retrospective cohort study using the Pediatric Heart Network (PHN) Single Ventricle Reconstruction trial public dataset, including 549 infants enrolled at 15 North American centers between 2005 and 2008. Competing risk analysis was used to evaluate Fontan completion and Cox regression to assess death or transplantation within 6 years after the Norwood procedure. Mixed-effects models compared pre-Fontan hemodynamic and echocardiographic right ventricular indices between patients treated with and without digoxin after accounting for center clustering and adjustment for sex, shunt type, heart failure medications at Norwood discharge, and census block poverty level

**Results:** The 6-year cumulative incidence of Fontan completion was higher among patients discharged on digoxin than among those not receiving digoxin (82% vs 71%; p = 0.013). Competing-risk analysis accounting for death and transplant demonstrated a greater likelihood of Fontan completion among digoxin users (aHR 1.31; 95%CI 1.09–1.58; p = 0.005), without significant difference in the hazard of death or transplant (aHR 0.78; 95%CI 0.53–1.15; p = 0.208). No significant differences in pre-Fontan hemodynamic or echocardiographic indices were observed between groups. Initiation of digoxin post Stage II procedure was not associated with improved survival or likelihood to complete Fontan.

**Conclusion:** Digoxin use at the time of Norwood discharge was associated with a 30% greater likelihood of Fontan completion by 6 years, without accompanying improvement in transplant-free survival. These findings extend prior observations of improved interstage outcomes associated with digoxin use and suggest that treatment may facilitate progression through staged palliation.

**Clinical perspective:** *What is new?:* Digoxin use during the interstage was associated with a greater likelihood of reaching Fontan completion within 6 years after Norwood discharge. However, this association was not accompanied by differences in right ventricular hemodynamic or echocardiographic indices at the time of Fontan evaluation.

*What are the clinical implications?:* These findings extend prior observations of improved interstage outcomes associated with digoxin use and provide important information for clinicians caring for patients with single-ventricle heart disease. Further studies are needed to clarify the mechanisms underlying this association and to identify patients most likely to benefit.

## Introduction

Patients with single ventricle physiology, particularly those undergoing the Norwood procedure, face a complex and challenging journey towards Fontan completion. The interstage period, spanning from the Norwood procedure to the Fontan operation, is a critical phase where medical interventions can have a profound impact on patient outcomes. In recent years, studies from the National Pediatric Cardiology Quality Improvement Collaborative (NPC-QIC) and the Pediatric Heart Network (PHN) have identified modifiable factors associated with improved interstage outcomes, findings that have been incorporated into the American Heart Association 2024 scientific statement. [1–5] .

Among these, digoxin use at discharge following the Norwood procedure has been consistently associated with reduced interstage mortality.[1–4, 6] In addition to its association with survival, digoxin use has been linked within the PHN cohort to preservation of echocardiographic indices of right ventricular (RV) performance, including RV end-systolic and diastolic area and ejection fraction during the interstage period. [7, 8] These measures of ventricular health have themselves been associated with improved survival, suggesting a potential mechanistic pathway through which digoxin may confer benefit. [9, 10] Despite these observations, it remains unclear whether the early advantages associated with digoxin use translate into improved progression through staged palliation or more favorable long-term outcomes.

We hypothesized that preservation of right ventricular health during the interstage period might influence both transplant-free survival and the ability to successfully progress to Fontan completion. Using data from the PHN/SVR (Single Ventricle Reconstruction) trial, we aimed to determine the association between digoxin use at Norwood discharge and both transplant-free survival as well as successful Fontan completion. We also assessed right ventricular hemodynamic and echocardiographic parameters at the time of Fontan evaluation according to digoxin use at Norwood discharge.

## Method

### Study Design

We performed a retrospective cohort study using data from the Pediatric Heart Network Single Ventricle Reconstruction Extension Study public dataset (downloaded from www.pediatricheartnetwork.org on 2/15/2023). The study was conducted across 15 North American centers, enrolling infants with single ventricle congenital heart disease and morphologically dominant RV, who underwent Norwood procedure between 2005 and 2008.[11–14] The main aim of the original study was to compare survival outcomes after procedures utilizing the Blalock-Thomas-Taussig shunt (BTTS) vs a RV to pulmonary artery conduit (RV-PA or Sano shunt).

The primary outcome in our study was successful completion of the Fontan pathway and transplant-free survival 6 years after the Norwood discharge, allowing for a uniform duration of follow up across the study cohort. Secondary outcomes included comparison of echocardiographic and hemodynamic parameters at the time of evaluation for the Fontan procedure between patients discharged on and without Digoxin following the Norwood procedure.

Subsequent operations [Stage II (Glenn) and Fontan procedures], as well as transplant and death events were obtained from the public dataset along with results of echocardiographic assessment at three prespecified time points (Norwood discharge, pre-Glenn, and pre-Fontan) as available based on the original trial protocol. Hemodynamic data were obtained from cardiac catheterization performed before the Glenn and Fontan procedures.

Institutional review board (IRB) approval and informed consent was obtained at participating institutions at the time of enrollment of the cohort. Since the public dataset contains no personal identifications, the present study was approved by Emory’s IRB with waiver of consent.

### Statistical Methods

Descriptive statistics were summarized as counts with percentages for categorical variables and as medians with interquartile ranges (IQR: Q1-Q3) for continuous variables. Group comparisons between patients discharged on Digoxin (yes vs. no) were performed separately at Norwood and Glenn discharge. Categorical variables were compared using Pearson’s Chi-squared test or Fisher’s exact test, as appropriate, while continuous variables were compared using the Wilcoxon rank-sum test.

Time to Fontan completion, between Digoxin groups, was evaluated using cumulative incidence functions (CIF) through 6 years of follow-up, treating death and heart transplant as competing events. Cumulative incidence estimates with 95% confidence intervals (95% CI) were reported at prespecified time points, and differences between groups were assessed using Gray’s test. Fine-Gray subdistribution hazard models were used to compare Fontan completion between Digoxin groups, with results reported as subdistribution hazard ratios (sHR) and 95% CI.

Transplant-free survival was assessed using Kaplan-Meier (KM) curves and Cox proportional hazard models through 6 years of follow-up. Differences between treatment groups were evaluated using the log-rank test. Cox models were reported as hazard ratios (HR) with corresponding 95% CI.

Multivariable Cox and Fine-Gray models adjusted for prespecified covariates selected based on clinical relevance and prior evidence of association with outcomes following the Norwood procedure, rather than solely on statistical differences between treatment groups. These included sex, shunt type, discharge on heart failure medications other than digoxin, and socioeconomic status as measured by census block poverty level. [11, 12, 15, 16] To account for center-level variation, site volume was included and categorized as ≥10 versus <10 Norwood procedures per year.[17, 18]

Sensitivity analyses included additional adjustment for arrhythmias and stratified analyses by arrhythmia presence. Adjusted models were reported as adjusted hazard ratios (aHR) or adjusted subdistribution hazard ratios (asHR), as appropriate, with 95% CIs.

Two exploratory analyses were conducted. First, to account for changes in exposure over time, digoxin use was classified based on exposure after the Norwood and Glenn procedures (Norwood only, Glenn only, both, or neither) and analyzed using the same time-to-event and competing risk approaches. Second, catheterization hemodynamics and echocardiographic right ventricular indices were analyzed using linear mixed-effects models to account for clustering by center and repeated measures within patients. A difference-in-differences (DiD) approach was used to evaluate changes between pre-Glenn to Fontan stage. Multivariable models were adjusted for shunt type, medication use, and age at Norwood. Least-squares means (LS-Means) and standard errors (SE) were reported and between-group differences are presented with 95% CIs. Effect sizes were quantified using Cohen’s *d* for comparisons of interest.

A two-sided p-value of < 0.05 was considered statistically significant. All analyses were conducted using SAS version 9.4 (SAS Institute Inc., Cary, NC) and R version 4.2.1 (R Foundation for Statistical Computing, Vienna, Austria).

## Results

### Study cohort and patient characteristics

A total of 549 infants undergoing the Norwood procedure between 2005 and 2008 met inclusion criteria. Of these, 437 survived to Norwood discharge (Figure 1). During the interstage period, 41 deaths, 6 heart transplants, and 1 loss to follow-up occurred in the group who did not receive Digoxin, while 11 deaths were reported in the Digoxin group, for a total of 52 deaths. Ultimately, 378 patients proceeded to Stage II (Glenn) surgery, with 10 additional in-hospital deaths, providing 368 patients who were discharged after Glenn.

**Figure 1.**
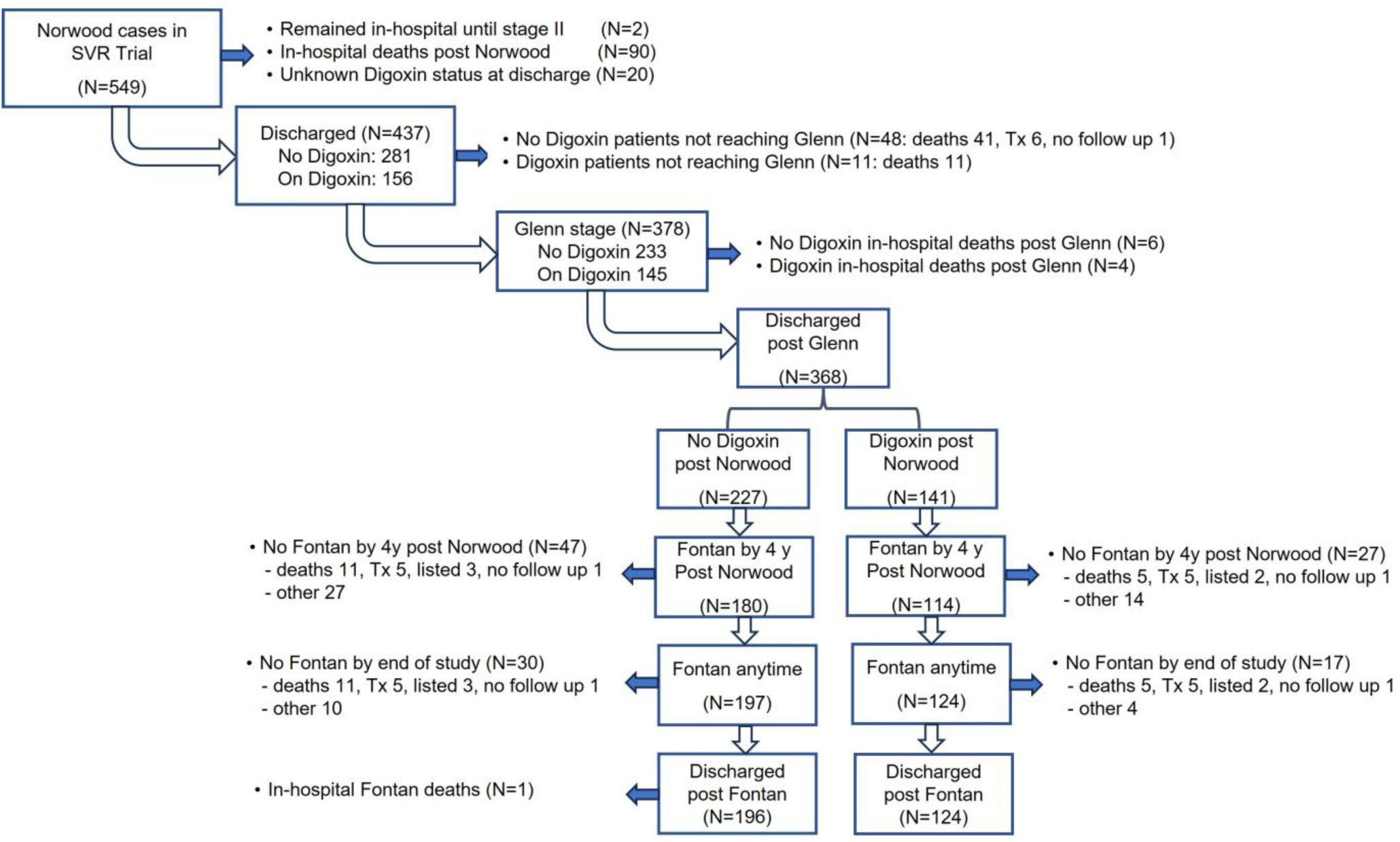
Patient flow diagram by digoxin exposure from Norwood to Fontan discharge.

By Glenn discharge (Supplemental Figure 1), treatment patterns had evolved: 46 patients initially discharged without digoxin were newly started on digoxin, while 90 patients initially discharged on digoxin had discontinued therapy. This resulted in four treatment subgroups for descriptive and secondary analyses, allowing evaluation of both interstage and post-Glenn digoxin exposure.

Baseline characteristics at Norwood discharge are summarized in Table 1. Patients discharged on digoxin (n = 156) and those not receiving digoxin (n = 281) were similar across most demographic, anatomic, and socioeconomic variables. The digoxin group had a higher prevalence of arrhythmia (35% vs. 22%, p = 0.003) and were less frequently discharged on other heart failure medications (82% vs. 93%, p < 0.001). No significant differences were observed in gestational age, birthweight, shunt type, or center surgical volume.

**Table 1.**
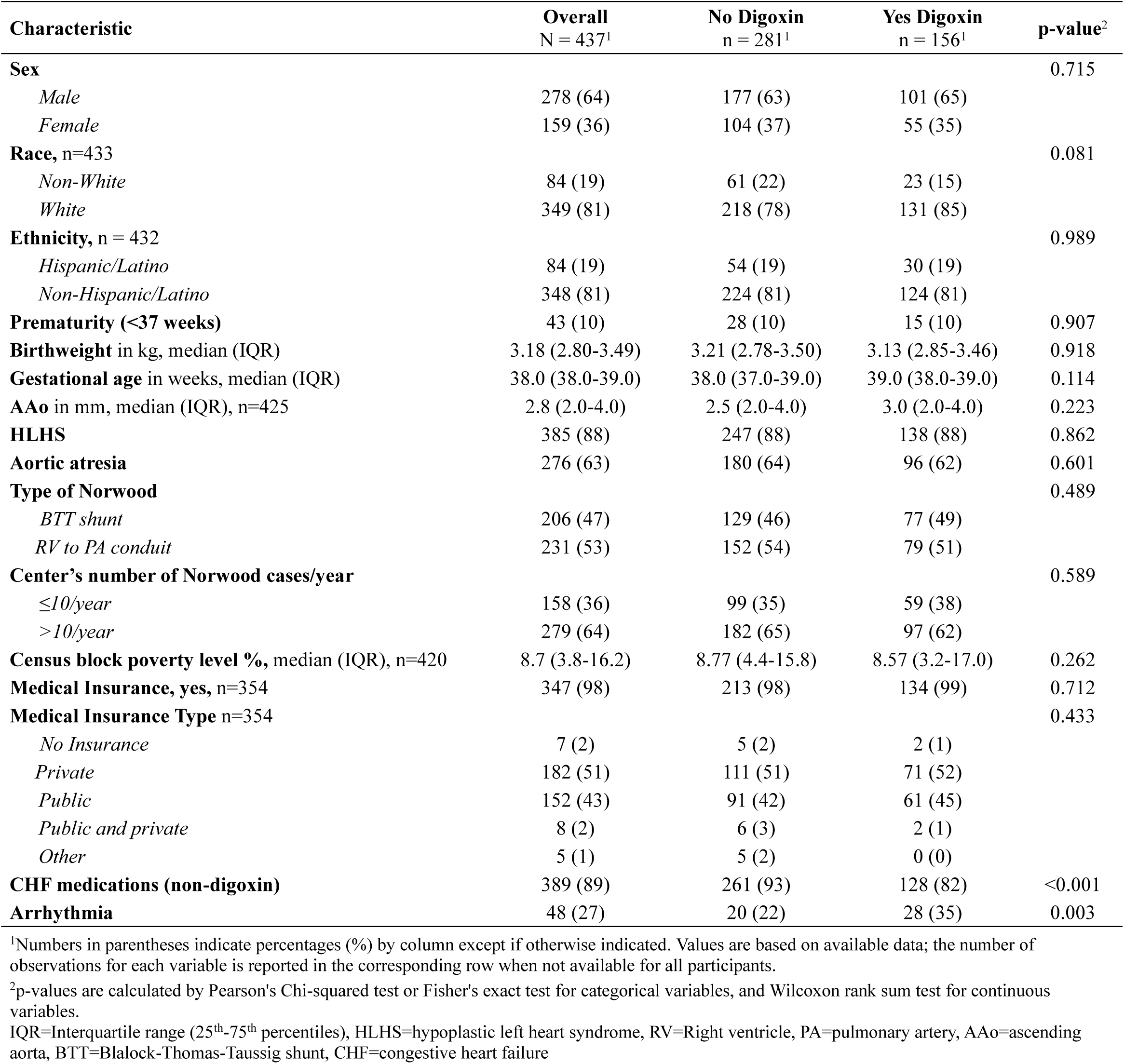
Patient and center characteristics by Digoxin treatment at Norwood discharge.

Among the 368 patients surviving to Glenn discharge and followed through Fontan (Table 2), demographic characteristics remained similar between groups. Patients discharged on digoxin were more likely to have hypoplastic left heart syndrome (98% vs. 85%, p < 0.001) and had smaller ascending aortic diameters (p=0.001). Shunt type, center volume, and socioeconomic indices did not differ. Fontan pathway type was similar between groups.

**Table 2.**
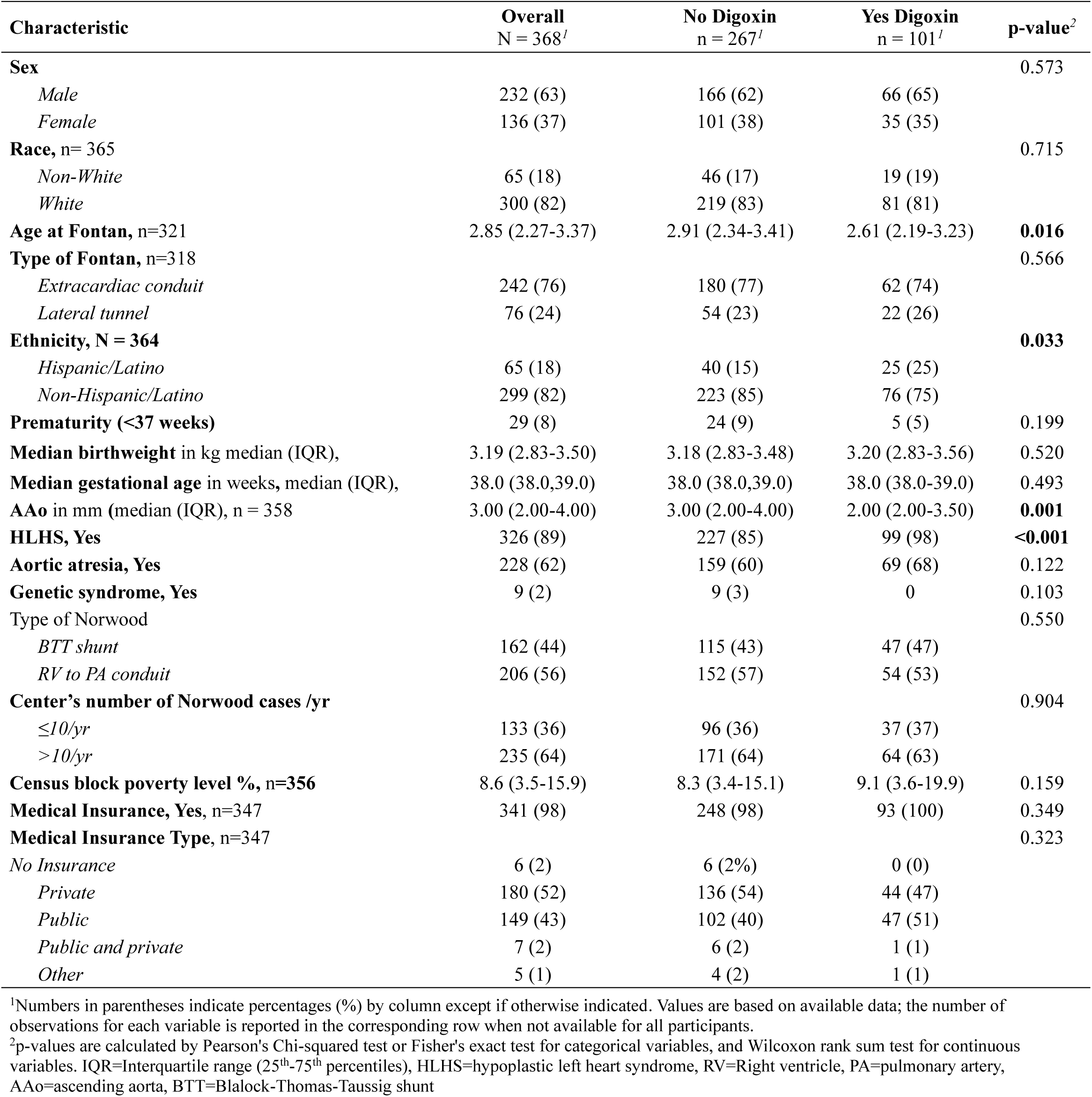
Characteristics of patients discharged at Glenn by Digoxin treatment at Norwood discharge.

### Fontan completion and transplant-free survival

Of the 368 patients who were discharged post Glenn, 321 (85%) completed Fontan palliation (124 in the digoxin group and 198 in the non-digoxin group). Most Fontan procedures (92%) occurred within 4 years of Norwood discharge, with a mean age of 2.9 years (IQR: 2.27-3.37) at Fontan completion.

The 6-year (median follow up time 2.52 yrs; IQR: 1.64-3.18) cumulative incidence of Fontan completion was higher among patients discharged on digoxin (82%; 95% CI 74–87%) compared with those not receiving digoxin (71%; 95% CI 66–76%; log-rank p = 0.013; Figure 2).

**Figure 2.**
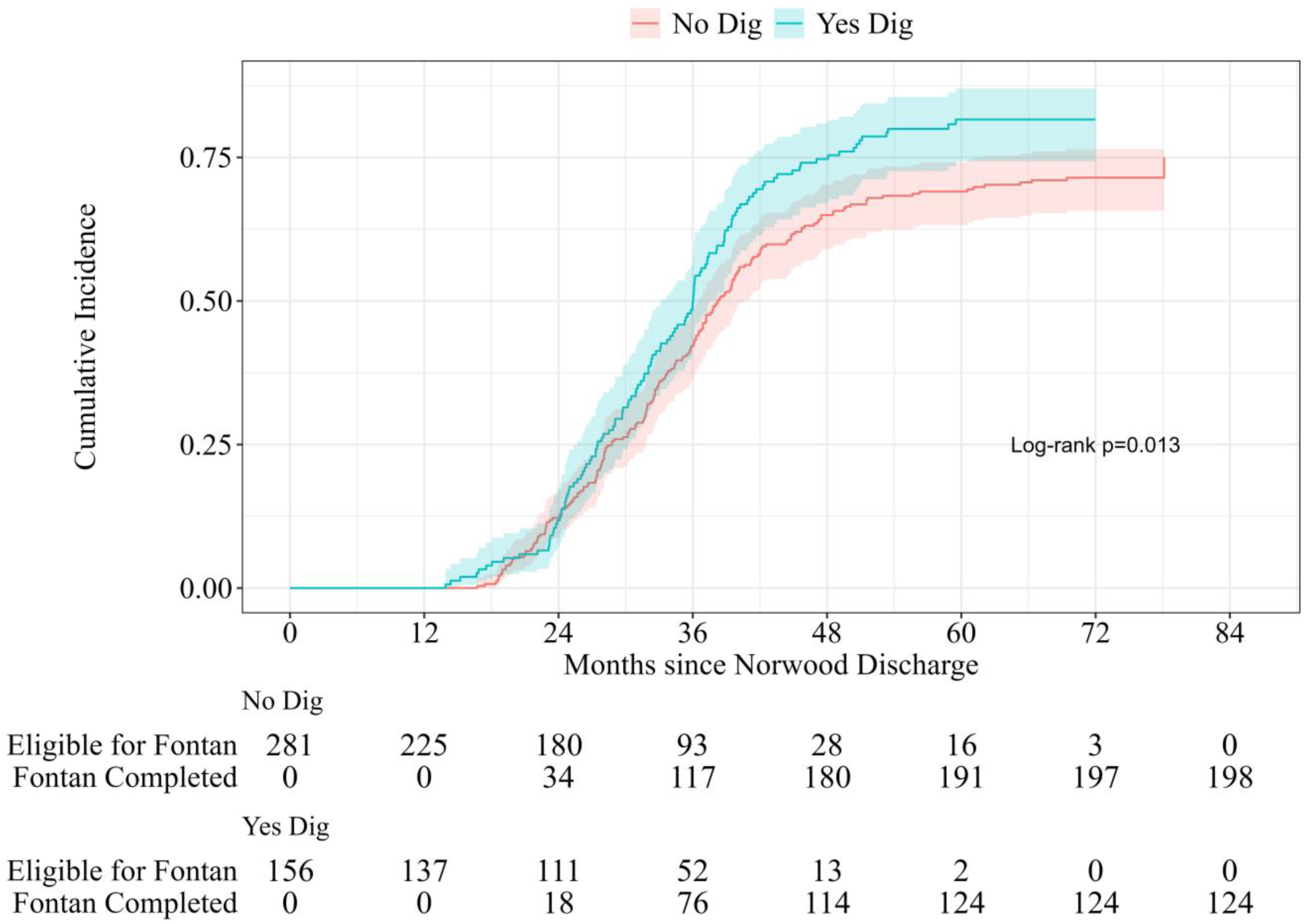
Cumulative incidence of Fontan completion among patients with and without Digoxin use at time of Norwood discharge through 6 years of follow up.

In competing-risk analyses accounting for death and transplant, digoxin use at Norwood discharge was associated with a higher likelihood of Fontan completion (sHR 1.31; 95% CI 1.09–1.58; p = 0.005) (Table 3). Univariable associations for all covariates are presented on Supplemental Table 1. This association persisted after multivariable adjustment for prespecified covariates (type of Norwood, use of CHF medications, sex and census poverty level) including arrhythmia (asHR 1.32; 95% CI 1.07–1.62; p = 0.01) (Table 3). In subgroup analyses, digoxin remained associated with a higher likelihood of Fontan completion among patients without arrhythmias (asHR 1.30; 95% CI 1.05–1.61; p = 0.017). A similar effect estimate was observed among patients with arrhythmias (asHR 1.35, 95% CI 0.85–2.14), although statistical significance was not reached, possibly reflecting the smaller sample size in this subgroup.

**Table 3.**
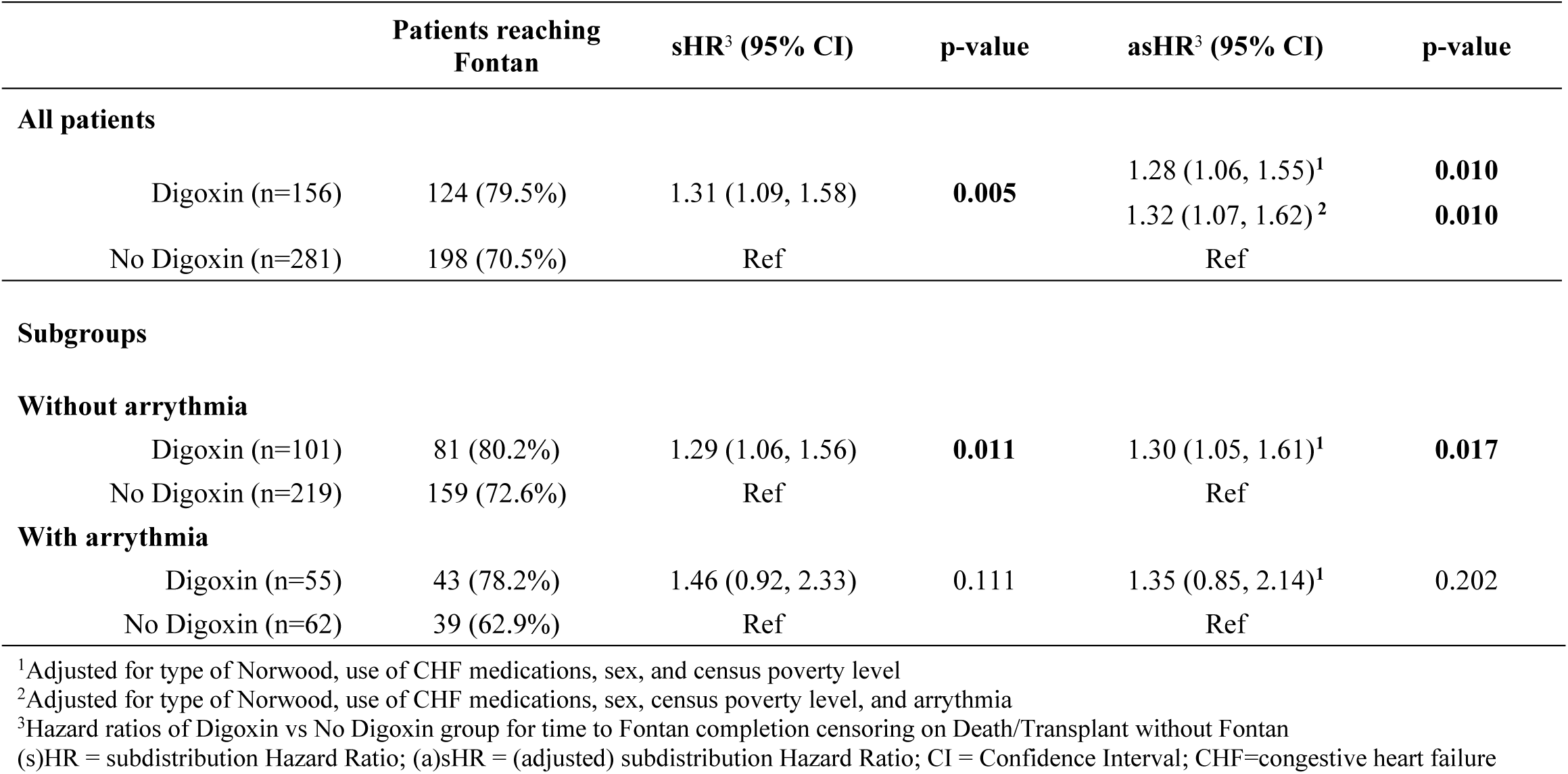
Analysis of Fontan completion by Digoxin treatment at Norwood discharge using Fine-Gray models.

Kaplan–Meier analyses demonstrated no statistically significant difference in transplant-free survival between digoxin and non-digoxin groups through 6 years following Norwood discharge (Figure 3). Adjusted Cox regression similarly showed no association between digoxin use at Norwood discharge and transplant-free survival (aHR 0.78; 95% CI 0.53–1.15; p = 0.208) within the observed follow-up period (Supplemental Table 2).

**Figure 3.**
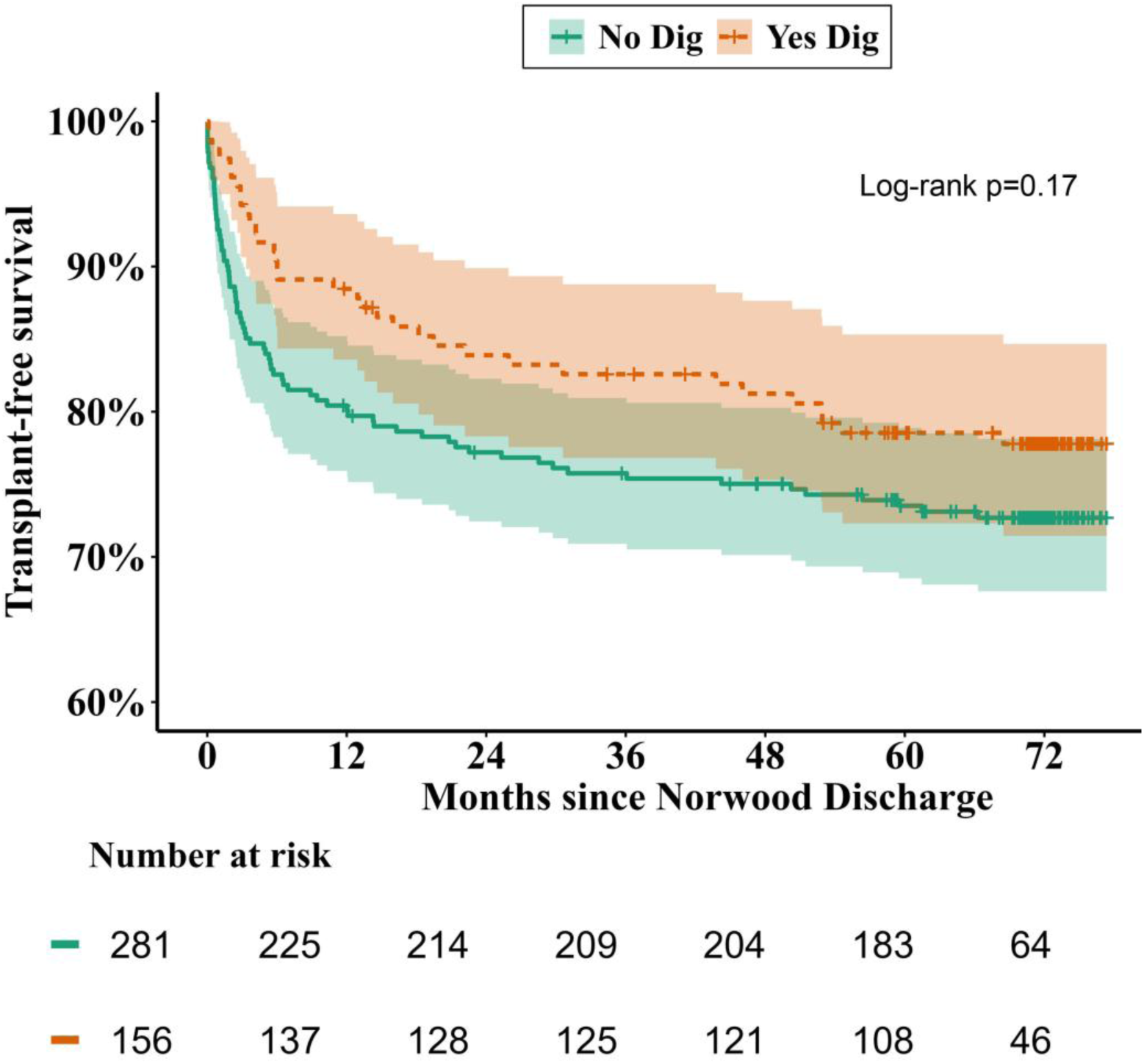
Kaplan-Meier transplant-free survival curves post Norwood by Digoxin treatment group from Norwood discharge through 6 years of follow-up.

Further stratification by digoxin use after the Glenn procedure distinguished 4 separate groups (Supplemental Figure 1). Among these four post-Glenn treatment subgroups, Fontan completion was not different between the 4 subgroups (Supplemental Figure 2 / Supplemental Table 3). However, the transplant-free survival was generally slightly lower among those treated with Digoxin either at Norwood or Glenn discharge (or both) (Supplemental Figure 3) with a corresponding higher observed hazard for post-Glenn death or transplant (Supplemental Table 4) for all subgroups except those with arrhythmias treated at Norwood discharge (+/- treated at Glenn discharge). After adjustment for the other covariates, statistically significant higher hazard remained only among patients without arrythmia who were newly initiated on digoxin after Glenn compared with those never treated with digoxin (aHR 2.55; 95% CI 1.06–6.10; p = 0.036).

### Hemodynamic and Echocardiographic Findings

There were no clinically or statistically significant differences in systemic ventricular end-diastolic pressure (SVEDP) or mixed venous oxygen saturation (SvO2) neither at the pre-Glenn nor pre-Fontan hemodynamic evaluations between those receiving and not receiving digoxin at Norwood discharge (Supplemental Table 5). Similarly, there were no differences between the right ventricular size and systolic function echocardiographic indices between groups (Supplemental Table 6).

## Discussion

In this multicenter cohort of infants with single ventricle physiology enrolled in the PHN/SVR trial, digoxin use at the time of Norwood discharge was independently associated with a significantly greater likelihood of Fontan completion by 6 years, without a corresponding improvement in long-term transplant-free survival. These findings extend prior observations of an interstage survival benefit associated with digoxin and suggest that early therapy may influence progression through staged palliation, even in the absence of a detectable long-term survival effect. While Fontan completion represents an important intermediate milestone, our findings indicate that improved progression to this stage does not necessarily translate into improved transplant-free survival within mid-term follow-up.

The observed association between digoxin use and Fontan completion appears to be driven primarily by an effect during the interstage period. Prior analyses from the Pediatric Heart Network and NPC-QIC have consistently demonstrated improved transplant-free survival among infants treated with digoxin during the vulnerable interstage period. [1–4, 7, 8] Digoxin’s benefit for transplant free survival was attributed to the preservation of echocardiographic right ventricular indices, including ventricular size and systolic function during the interstage period, [8,9] which had been independently associated with improved survival and downstream outcomes in single ventricle patients. [10]

In the present study examining post Glenn follow-up data from the same cohort, digoxin-treated patients demonstrated non-significant differences in the hemodynamic data (SVEDP and SvO_2_) at the pre-Fontan cardiac catheterization, while the preservation of the RV echocardiographic indices was no longer present by the time of Fontan evaluation. These findings suggest that any beneficial effects of digoxin may be transient and related to early clinical stabilization of a likely high-risk group during the interstage period, which is consistent with prior reports on that subject [2]. Although digoxin use was associated with a greater likelihood of progressing through staged palliation, this association was not accompanied by persistent differences in ventricular performance or hemodynamic measures at the time of Fontan evaluation. These findings suggest that any benefit of digoxin may be mediated through mechanisms operating during the interstage period rather than through sustained improvements in ventricular remodeling. Digoxin initiation after Stage II (Glenn) was not associated with improved survival or Fontan completion. The observed increase in post-Glenn hazard for death or transplant among patients newly started on therapy after Glenn is most likely explained by confounding by indication, whereby initiation of therapy reflects worsening clinical status rather than a harmful effect of digoxin itself. The association of digoxin use in patients with high-risk clinical features such as HLHS, small ascending aortas and arrhythmias, further supports this interpretation.

As a retrospective observational analysis, this study is subject to non-random treatment allocation. Selection bias and confounding by indication are likely present, as clinicians may have preferentially prescribed digoxin to infants perceived to be at higher risk, including those with ventricular dysfunction or early heart failure symptoms. In particular, late initiation of digoxin after Glenn appears to identify a clinically deteriorating subgroup, in which case digoxin’s use is a marker of worsening clinical status rather than a causal contributor to adverse outcomes.

Concerns regarding digoxin toxicity and pro-arrhythmic potential in congenital heart disease populations have been raised in prior editorial commentary, underscoring the importance of careful patient selection and monitoring rather than routine use.[19] However, these concerns have not been accompanied by evidence demonstrating net harm when digoxin is used during the interstage period. In contrast, multiple observational analyses from PHN and NPC-QIC consistently demonstrate improved outcomes with interstage digoxin therapy. [1–3, 20, 21] In the absence of randomized trial data, clinicians must balance recognized risks against the best available observational evidence and the practical challenges of conducting randomized trials in this population. Contemporary pediatric heart failure reviews continue to acknowledge digoxin as a therapy with a limited but potentially meaningful role in selected congenital heart disease populations while highlighting persistent evidence gaps and practice variability.[20, 22–24]

Despite ongoing clinical equipoise, randomized controlled trials evaluating digoxin in interstage single-ventricle patients are unlikely to take place due to ethical concerns, small population size, and entrenched clinician preferences. Accordingly, large, multicenter observational datasets remain essential for informing clinical practice in this setting. Further studies are needed to clarify the mechanisms underlying digoxin’s beneficial effects during interstage and to identify patients most likely to benefit.

### Limitations

Several limitations warrant consideration. This study represents a secondary analysis of the PHN/SVR public dataset, and digoxin was not prescribed randomly. Thus, we are unable to determine the specific clinical rationale for initiating, continuing, or discontinuing digoxin, limiting our ability to distinguish treatment effects from clinician-driven risk stratification. Prescription practices varied across centers and over time, and residual confounding cannot be fully excluded. Survivorship bias is inherent, as only patients surviving to Norwood discharge were included. Detailed data on post-discharge adherence, dosing, duration of therapy, and timing of discontinuation were unavailable. Arrhythmia diagnoses were site-reported and subject to misclassification. Finally, unmeasured anatomic or socioeconomic factors may have influenced outcomes despite multivariable adjustment.

### Conclusions

In summary, digoxin use at the time of Norwood discharge was associated with approximately a 30% greater likelihood of Fontan completion by 6 years, without an accompanying improvement in long-term transplant-free survival. These findings support the continued use of digoxin during the interstage period in appropriately selected patients, where it may facilitate stabilization and progression through staged palliation. In contrast, no evidence of improved survival or progression to Fontan was observed among patients receiving digoxin after the Glenn procedure, although these findings should be interpreted cautiously given the likelihood of confounding by indication.

## Data Availability

The data are publicly available as part of the Pediatric Heart Network Single Ventricle Reconstruction Extension Study public dataset (downloadable from). SAS codes can be available by the corresponding author upon reasonable request.

https://www.pediatricheartnetwork.org

